# The association between street construction projects and community violence in New York City

**DOI:** 10.1101/2024.05.30.24308120

**Authors:** Brady Bushover, Andrew Kim, Christina A. Mehranbod, Leah E. Roberts, Ariana N. Gobaud, Evan L. Eschliman, Carolyn Fish, Xiang Gao, Siddhesh Zadey, Christopher N. Morrison

**Author notes:** **Correspondence to:** Christopher N. Morrison, Department of Epidemiology, Columbia University Mailman School of Public Health, 722 W 168^th^ St, Rm 505, New York, NY 10032.

## Abstract

**Introduction:** Community violence is a major cause of injury and death in the United States. Empirical studies have identified that some place-based interventions of urban private places, such as remediations of vacant lots and buildings, are associated with reductions in community violence in surrounding areas. The aim of this study was to examine whether routine maintenance and repair of urban public places (e.g., street construction projects) are also associated with reductions in community violence, proxied by violent crime.

**Method:** This staggered adoption difference-in-difference analysis investigated the association between street construction projects and community violence in New York City from 2010-2019, divided into 40 calendar quarters. The units of analysis were street-quarters (n = 155,280). Intervention street-quarters were those with completed projects in 2010-2019; control streets were those where projects were scheduled but not completed before 2019. The outcome of community violence was proxied by counts of crime and violence incidents reported to the New York Police Department, within street-quarters.

**Results:** There were 79,592 street-quarters with any community violence incidents (51.2%). We found that street construction projects were associated with a decrease in reckless endangerment (ATT = -0.013; 95% CI = -0.021, -0.004), robbery (ATT = -0.035; 95% CI = -0.063, -0.007), and weapons offenses (ATT = -0.016; 95% CI = -0.031, -0.001) occurring on street-quarters.

**Conclusion:** Street construction projects may be yet another type of place-based intervention to reduce community violence.

## Introduction

Community violence, defined as “the intentional use of physical force or power, threatened or actual, against another person, group or community in a specific location that results in or has a high likelihood of resulting in injury, death, psychological harm, maldevelopment, or deprivation,” ^1^ is a major cause of injury and death in the United States (US). Homicide, one type of community violence, is a leading cause of death for those aged one to 44. ^2^ In 2022, nearly 25,000 people died due to homicide, and over 300,000 were hospitalized due to a violence-related injury.^2^ In addition to injuries and death, community violence represents a large economic burden. When looking at community violence through the lens of violent crime, violent crime is estimated to result in a $2 trillion cost in monetary and quality of life losses each year.^3^ Interventions at the population level provide upstream opportunities to reduce these preventable deaths, injuries, and associated economic costs.^4^

One such type of population-level intervention to reduce community violence involves altering the physical environments at places where violence concentrates. Previous studies have documented associations between place-based interventions and decreases in reports of violent crime. ^5^ These modifications to the built environment mostly include interventions implemented in privately owned places, such as greening vacant lots^6^ and demolishing vacant buildings.^7^ For example, a citywide cluster randomized trial in Philadelphia, Pennsylvania found that vacant lots that received a greening treatment were associated with 8.7% reduction in all police-reported crime compared to control lots.^8^ Observational studies conducted in various locations across the US support the findings of experimental studies.^9-11^ Community-engaged vacant lot maintenance was associated with declines in several different types of crimes, including assault and robbery, in Youngstown, Ohio.^12^ And in Milwaukee, Wisconsin the conversion of vacant lots into community gardens was associated with a 3.7-6.4% reduction in violent crime.^13^

Several theoretical mechanisms could explain how place-based interventions affect community violence. “Busy streets” theory^14^ suggests that removing signs of physical disorder in a neighborhood creates opportunities for positive social interactions, reduces fear and pessimism, and increases perceptions of safety among neighborhood residents. For example, participants living near treated vacant lots in the aforementioned randomized trial of place-based interventions in Philadelphia reported a 60% reduction in fear of going outside because of safety concerns after remediations were completed. ^8^ Complementarily, features of the built environment have been shown to be positively associated with collective efficacy,^15^ which is the social cohesion of a neighborhood and residents’ willingness to intervene on behalf of the common good.^16^ Place-based interventions may increase the number and frequency of social interactions—as both a form of active surveillance and a contributor to collective efficacy—and lead to improvements in a variety of intrapersonal factors, ultimately inhibiting community violence.^17^

It follows that major public works projects may also lead to reductions in community violence, even if that is not their primary function. While previous work on place-based interventions has focused on private places like buildings and vacant lots, altering public roadways may also influence individuals’ feelings and perceptions related to their neighborhood. Projects such as resurfacing streets or repairing sidewalks could promote social interactions among residents. Both mechanisms may reduce community violence. Previous research has shown that place-based interventions targeting the street environment specifically are associated with reductions in crime and violence. For example, an experimental study in New York City found that street lighting at night decreased violent crime.^18^ However, no studies have investigated the association between community violence and street construction projects.

The aim of this study was to conduct a natural experiment using highly resolved space-time data for street construction projects and community violence—measured by reports of violent crime—in New York City. We hypothesized that street segments that received a street construction project would have lower incidence of community violence compared to street segments that did not receive any construction projects.

## Methods

### Ethics

This study utilized only publicly available data and was determined not to be human subjects research by the Columbia University Institutional Review Board.

### Study sample

We used a staggered difference-in-difference analysis to assess the relationship between road construction projects and community violence. The study period included 10 years from January 1, 2010, to December 31, 2019, divided into 40 calendar quarters. The units of analysis were street-quarters. Street segments that were assigned to receive a street construction project between January 1, 2010, and December 31, 2019, were eligible for inclusion. Intervention street-quarters were those with completed projects. Control street-quarters were those with scheduled projects that were canceled or did not have construction activities start until after December 31, 2019. Using this inclusion criterion allowed us to avoid positivity violations^19^ and ensure that intervention and control streets were comparable in that each included street was identified as eligible to receive a street construction project by the New York City Department of Transportation (i.e., the agency responsible for the coordination of all street construction projects).

### Data

The dependent measure of interest was community violence, which we operationalized as violent crimes using a registry of crimes reported to the New York Police Department (NYPD).^20^ Crime data are geocoded by the NYPD and coordinates for the location of each incident are provided in the publicly available dataset. We extracted unique incidents by crime type that occurred between January 1, 2010, and December 31, 2019, and matched them to the street-quarter on which they took place. We then summed the occurrence of each incident type for each street-quarter. Violent crime types were based on New York Penal Law^21^ and included: felony assault, grand theft auto, murder, reckless endangerment, robbery, and weapons offenses. Reckless endangerment involves acting in a way that creates a risk of serious physical injury or death for someone else, and weapons offenses include criminal possession or sale of a weapon.

The main independent variable was the completion of a street construction project. The New York City Department of Transportation provides publicly available, geocoded data for street construction projects at the street segment level.^22^ We obtained data for all projects that occurred on street segments between January 1, 2010, to December 31, 2019. Street-quarters were assigned a variable to indicate intervention status. Once a street segment was assigned as having received the treatment, it was considered to have remained exposed to the treatment for the rest of the study period.

There are several different types of street construction projects in New York City, varying in scope (Fig. 1). For example, street resurfacing replaces the top layer of pavement on a roadway and typically requires a month to complete, while street reconstruction involves the replacement of over a foot of the roadway beneath the ground and will commonly include the replacement of curbs and/or sidewalks. The completion of street reconstruction projects may take several years.^23,24^ Other types of construction projects include the repair or installation of bike lanes, bus lanes, light posts, and pedestrian ramps. Any type of construction project that affects the roadbed, such as sewer system repairs, are also included as street construction projects. Projects are initiated based on demonstrated need and community input, and can occur at any time.^24^

**Fig. 1.**
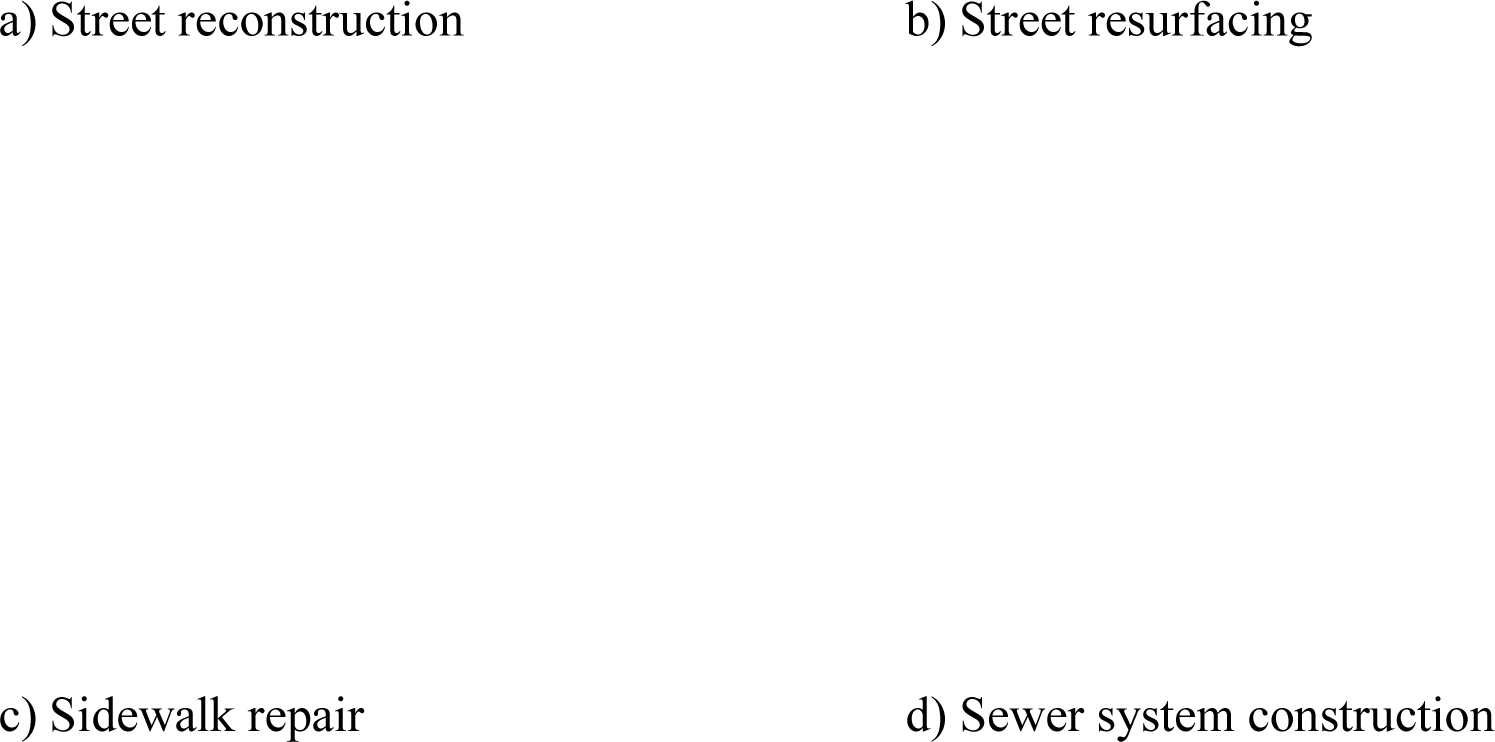
Examples of street construction project types. Images of a) street reconstruction, b) street resurfacing, c) sidewalk repair, and d) sewer system projects in New York City.^30^ Please contact the corresponding author to request access to these images.

### Statistical analysis

For this difference-in-difference analysis, group-time average treatment effect estimators were calculated for the association between street construction projects and community violence. Since the street construction project intervention was not implemented citywide at a single timepoint, a staggered difference-in-difference design was required. This design allowed us to account for the varying times of street construction project completion across all included street-quarters and comparing within-street effects. While fixed effects models are often used with panel data for difference-in-difference estimations, they are not robust to heterogenous effects and multiple time periods.^25,26^ Thus, we used the staggered adoption difference-in-difference approach developed by Callaway and Sant’Anna^27^ to calculate average treatment effect on the treated (ATT) estimates for each crime type.

The ATT for each quarter in which at least one street segment received the treatment was estimated. Each quarter in which a treatment was completed comprises an adoption cohort. Once every ATT for each quarter where a treatment occurred was estimated, the ATTs were summarized into an overall estimate of the effect of street construction projects on community violence, and the effect estimate for each adoption cohort was produced. This group-time average treatment effect is calculated as:

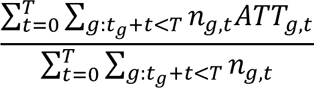

Where *n_g,t_* is the number of street segments in adoption cohort *g* treated in quarter *t*. *ATT_g,t_* is the estimate for the ATT of adoption cohort *g* in quarter *t*. The denominator corresponds to the total number of treated street-quarter points.

Analyses were completed using R 4.3.2 (Vienna, Austria) and ArcGIS Pro 3.1.0 (Redlands, California).

## Results

Table 1 presents descriptive statistics of included street-quarters. Out of the 155,280 included street-quarters, 80,280 (51.7%) ever received a street construction project. There were 79,572 (51.2%) street-quarters that experienced any occurrence of community violence as measured via violent crime reports. By violent crime type, the proportion of street-quarters experiencing any violent crimes during the study period ranged from 0.2 (murder) to 14.8% (robbery). The most common project type was street reconstruction (86.3%). Projects took place across the entire extent of New York City, with most projects taking place in Brooklyn (40.4%). The locations of street segments with construction projects during the study period are presented in Fig. 2.

**Fig. 2.**
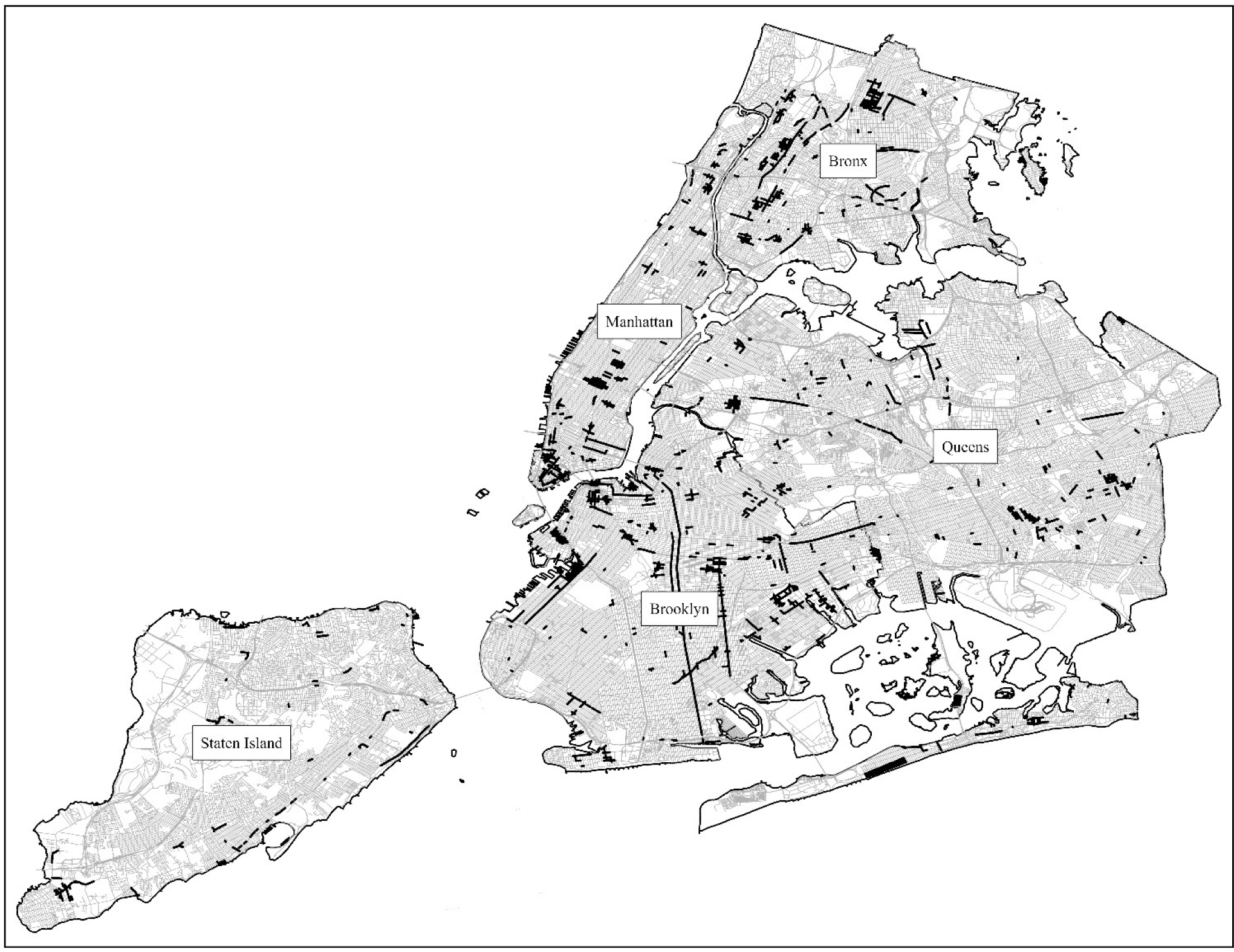
Street construction project locations in New York City, 2010-2019. Map of street segments with street construction projects overlayed on New York City streets, 2010-2019.

**Table 1.** Descriptive statistics for included street-quarters, 2010-2019.

|  |  |  |
| --- | --- | --- |
| N = 155,280 | **n** | **%** |
| Treated | 80,280 | 51.7 |
| Violent crime (ever experienced) |  |  |
| *Assault* | 19,068 | 12.3 |
| *Grand theft auto* | 6,044 | 3.9 |
| *Murder* | 265 | 0.2 |
| *Reckless endangerment* | 2,543 | 1.6 |
| *Robbery* | 22,909 | 14.8 |
| *Weapons offenses* | 8,678 | 5.6 |
| Project types |  |  |
| *Bus lane* | 6,987 | 4.5 |
| *Facilities* | 1,397 | 0.9 |
| *Highway repair* | 1,709 | 1.1 |
| *Lighting* | 155 | 0.1 |
| *Pedestrian ramps* | 932 | 0.6 |
| *Sewer* | 1,863 | 1.2 |
| *Sidewalk repair* | 932 | 0.6 |
| *Street reconstruction* | 134,007 | 86.3 |
| *Street resurfacing* | 7,298 | 4.7 |
| Borough |  |  |
| *The Bronx* | 24,224 | 15.6 |
| *Brooklyn* | 62,733 | 40.4 |
| *Manhattan* | 25,621 | 16.5 |
| *Queens* | 34,161 | 22.0 |
| *Staten Island* | 8,540 | 5.5 |

Table 2 shows the results of the staggered adoption difference-in-difference models. The confidence intervals for the aggregated ATTs for assault (ATT = -0.015; 95% CI = -0.041, 0.012), grand theft auto (ATT = 0.004; 95% CI = -0.006, 0.013), and murder (ATT = -0.003; 95% CI = -0.006, 0.000) crossed the null value. The completion of street construction projects were negatively associated with reckless endangerment (ATT = -0.013; 95% CI = -0.021, - 0.004), robbery (ATT = -0.035; 95% CI = -0.063, -0.007), and weapons offenses (ATT = -0.016; 95% CI = -0.031, -0.001).

**Table 2.** Average effect estimates across street-quarters by crime type.

| **Violent crime** | **Group-time average**<br>**ATT** | **95% CI** |
| --- | --- | --- |
| Assault | -0.015 | -0.041, 0.012 |
| Grand theft auto | 0.004 | -0.006, 0.013 |
| Murder | -0.003 | -0.006, 0.000 |
| Reckless endangerment | **-0.013** | **-0.021, -0.004** |
| Robbery | **-0.035** | **-0.063, -0.007** |
| Weapons offenses | **-0.016** | **-0.031, -0.001** |

Bolded values are statistically significant at an $\alpha$ of 0.05.

Fig. 3 shows the estimates of group-time average treatment effects by length of exposure, representing the average effect across adoption cohorts by the number of quarters either before or after the intervention, with a simultaneous confidence interval. The three violent crime outcomes for which a significant overall effect was found are included: reckless endangerment, robbery, and weapons offenses.

**Fig. 3.**
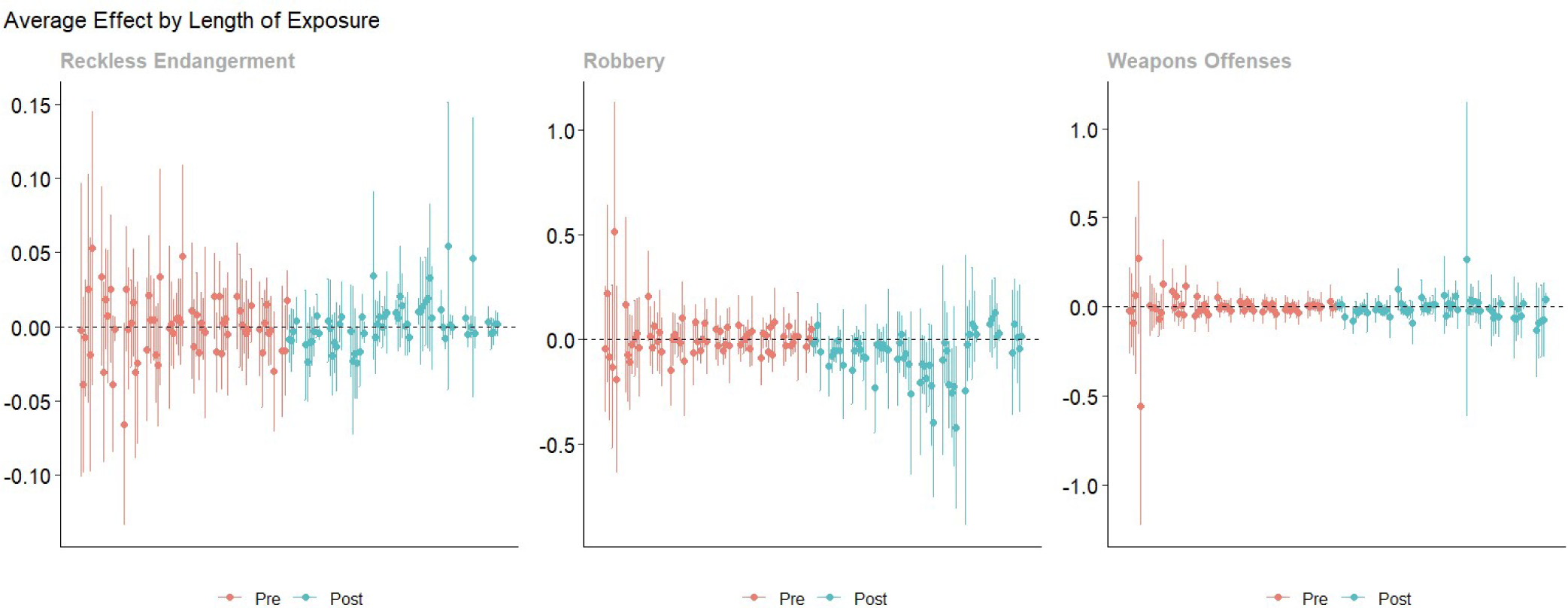
Group-time average treatment effect estimators by length of exposure. Estimates by length of exposure for the effect of street construction projects on reckless endangerment, robbery, and weapons offenses.

## Discussion

This study of street construction projects and community violence found that street segments that received a street construction project experienced decreases in several types of violent crimes as compared to street segments that did not. Our results indicate that street construction projects may be an effective place-based intervention to prevent community violence, with an estimated 1.3-3.5% decrease in incidents of reckless endangerment, robbery, and weapons offenses for streets that received a construction project.

Theoretical mechanisms, such as the busy streets theory and collective efficacy, that connect other place-based interventions to community violence may extend to street construction projects. These projects may be acting on interpersonal and intrapersonal factors in similar ways as treatments to vacant lots and vacant buildings. Furthermore, while many if not most greening and building remediation projects focus on interventions to private property, street construction projects act on public property. This type of intervention could thus provide local governments with a violence reduction strategy that concurrently improves public infrastructure. Previous work has shown that vacant building remediation and vacant lot greening, in addition to reducing community violence, is cost beneficial. For example, a study in Philadelphia, Pennsylvania found that for every dollar spent on vacant building remediation, the return to taxpayers as a result of firearm assaults averted was $5. For vacant lot remediations, it was $26.^28^ Since street construction projects are also associated with reductions in violence in New York City, they may present a secondary benefit to the routine maintenance and repair of public places.

These findings build on prior work demonstrating the value of place-based interventions in reducing community violence. While there are no previous studies testing the effects of public works projects on violence in New York City, a study assessing the use of nighttime street lighting found that index crimes were reduced by 35%.^18^ Studies in other cities have found reductions of violence and crimes after the implementation of place-based interventions. In Flint, MI, community-engaged greening of vacant lots was associated with an almost 40% reduction in assaults and total violent crime as compared to vacant lots not maintained by the community.^11^ Demolitions of structurally dangerous buildings in Saginaw, MI were associated with a 3.2% reduction in violent crimes and a 9.7% reduction in crime overall.^9^ Our estimates for the effect of street construction projects on community violence also show a negative association. While the magnitude of the association differs from previous work, our analysis incorporated a different space-time scale.

Assessing the effect estimates by length of exposure shows that the effects of street construction projects on community violence may decay over time. The average ATTs for greater lengths of time exposed exhibit weaker negative associations as compared to shorter ones, especially for reckless endangerment and robbery. This may present modest evidence of a decline in efficacy over time. It could also suggest that an effect is seen during the construction period. There may be protective effects of having construction workers present on the street. However, this pattern is not seen with weapons offenses. While a negative association was found for all three of these crime types over the ten years in this analysis, the efficacy of this intervention may differ among crimes for smaller periods of time. Determining the cause for this decay in effect will be important for developing successful street construction interventions aimed at violence prevention.

Examining the spatial and temporal decay of these effects will be important for designing effective interventions. In this study, we only assessed individual street segments over calendar quarters and did not account for spillover effects. Theoretically, the effects of road construction projects could extend to adjacent street segments. Also, street construction projects are long lasting. For example, once a street has been resurfaced or reconstructed in New York City it becomes a “protected street” and is ineligible for major street construction projects for five years.^29^ With a long-term intervention like this, decay of the observed effect could be gradual.

### Limitations

There are important limitations of this study to note. We only assessed the effects of construction projects with a post-treatment period. It is possible that the treatment had effects during the construction process. As a natural experiment, there may be sources of unknown confounding that we were unable to measure. Only streets that were scheduled to receive a street construction project were included. There may be systematic differences between streets that are selected to receive a project and those that are not in regard to crime incidents. Furthermore, reports of violent crime likely do not capture all of the incidents of community violence in a given area, yet they provide the best available estimates. Finally, this study investigates effects in one city. Other local areas may not experience the same effects as seen in New York City. Despite these constraints, our analysis benefitted from highly resolved space-time data on both street construction projects and incidents of community violence.

## Conclusions

This study adds to the body of literature supporting the use of place-based interventions to reduce community violence. Street construction projects may provide an additional tool for curbing community violence in a way that is congruent with the mechanisms by which already-proven approaches function. As community violence continues to impact communities across the US, it is critical to investigate and adjust effective interventions and reduce its burden. Future work is needed to examine the effects of street construction projects over time and across different cities.

## Data Availability

All data are available online at: https://opendata.cityofnewyork.us/

## Acknowledgements

**Funding and Support**: This work was supported with funding provided by grant R49CE003094 from the National Center for Injury Prevention and Control of the Centers for Disease Control and Prevention. ELE and ANG are supported by the National Institutes of Health, National Institute on Drug Abuse (T32DA031099). The findings and conclusions in this paper are those of the authors and do not necessarily represent the views of the funders.

**Role of the Funder/Sponsor**: The funders had no role in the design and conduct of the study; collection, management, analysis, and interpretation of the data; preparation, review, or approval of the manuscript; and decision to submit the manuscript for publication.

**Data Sharing Statement**: All data used for this study are publicly available. The dataset used in this study may be made available upon reasonable request to the corresponding author.

## Notes

### Competing Interest Statement

The authors have declared no competing interest.

## References

1. National Criminal Justice Association. An Overview: Community Violence Intervention Strategies. 2021. https://www.ncja.org/_files/ugd/cda224_c5b96183fb614e9692f99513646abd0d.pdf?index=true

2. Centers for Disease Control and Prevention. CDC Web-based Injury Statistics Query and Reporting System (WISQARS). Accessed May 13, 2024, https://www.cdc.gov/injury/wisqars/index.html

3. Miller TR, Cohen MA, Swedler DI, Ali B, Hendrie DV. Incidence and costs of personal and property crimes in the USA, 2017. *J Benefit Cost Anal*. 2021;12(1):24-54. doi:10.1017/bca.2020.36

4. Smith LS, Wilkins NJ, McClure RJ. A systemic approach to achieving population-level impact in injury and violence prevention. Syst Res Behav Sci. 2021;38(1):21–30. doi:10.1002/sres.2668

5. Gobaud AN, Jacobowitz AL, Mehranbod CA, Sprague NL, Branas CC, Morrison CN. Place-based interventions and the epidemiology of violence prevention. Curr Epidemiol Rep. 2022;9(4):316–325. doi:10.1007/s40471-022-00301-z

6. Moyer R, MacDonald JM, Ridgeway G, Branas CC. Effect of remediating blighted vacant land on shootings: A citywide cluster randomized trial. Am J Public Health. 2019;109(1):140–144. doi:10.2105/ajph.2018.304752

7. Jay J, Miratrix LW, Branas CC, Zimmerman MA, Hemenway D. Urban building demolitions, firearm violence and drug crime. J Behav Med. 2019;42(4):626–634. doi:10.1007/s10865-019-00031-6

8. Branas CC, South E, Kondo MC, et al. Citywide cluster randomized trial to restore blighted vacant land and its effects on violence, crime, and fear. Proc Natl Acad Sci U S A. 2018;115(12):2946–2951. doi:10.1073/pnas.1718503115

9. Stacy CP. The effect of vacant building demolitions on crime under depopulation. J Reg Sci. 2018;58(1):100–115. doi:10.1111/jors.12350

10. South EC, MacDonald J, Reina V. Association between structural housing repairs for low-income homeowners and neighborhood crime. JAMA Netw Open. 2021;4(7):e2117067. doi:10.1001/jamanetworkopen.2021.17067

11. Heinze JE, Krusky-Morey A, Vagi KJ, et al. Busy streets theory: The effects of community-engaged greening on violence. Am J Community Psychol. 2018;62(1-2):101–109. doi:10.1002/ajcp.12270

12. Kondo M, Hohl B, Han S, Branas C. Effects of greening and community reuse of vacant lots on crime. Urban Stud. 2016;53(15):3279–3295. doi:10.1177/0042098015608058

13. Beam DR, Szabo A, Olson J, Hoffman L, Beyer KMM. Vacant lot to community garden conversion and crime in Milwaukee: A difference-in-differences analysis. Inj Prev. 2021;27(5):403–408. doi:10.1136/injuryprev-2020-043767

14. Aiyer SM, Zimmerman MA, Morrel-Samuels S, Reischl TM. From broken windows to busy streets: A community empowerment perspective. Health Educ Behav. 2015;42(2):137–147. doi:10.1177/1090198114558590

15. Cohen DA, Inagami S, Finch B. The built environment and collective efficacy. Health Place. 2008;14(2):198–208. doi:10.1016/j.healthplace.2007.06.001

16. Sampson RJ, Raudenbush SW, Earls F. Neighborhoods and violent crime: A multilevel study of collective efficacy. Science. 1997;277(5328):918-924. doi:10.1126/science.277.5328.918

17. Cullen F, Wilcox P. Encyclopedia of Criminological Theory. SAGE Publications, Inc.; 2010. https://sk.sagepub.com/reference/criminologicaltheory

18. Chalfin A, Hansen B, Lerner J, Parker L. Reducing crime through environmental design: Evidence from a randomized experiment of street lighting in New York City. J Quant Criminol. 2022;38(1):127–157. doi:10.1007/s10940-020-09490-6

19. Petersen ML, Porter KE, Gruber S, Wang Y, van der Laan MJ. Diagnosing and responding to violations in the positivity assumption. Stat Methods Med Res. 2012;21(1):31–54. doi:10.1177/0962280210386207

20. NYC Open Data. Data from: NYPD Complaint Data Historic. 2024. https://data.cityofnewyork.us/Public-Safety/NYPD-Complaint-Data-Historic/qgea-i56i/about_data

21. Chapter 40: Penal. The New York State Senate. Updated September 22, 2014. Accessed May 17, 2024, https://www.nysenate.gov/legislation/laws/PEN/P3

22. NYC Open Data. Data from: Street and Highway Capital Reconstruction Projects - Block. 2023. https://data.cityofnewyork.us/Transportation/Street-and-Highway-Capital-Reconstruction-Projects/jvk9-k4re/about_data

23. New York City Department of Transportation. Street and Roadway Construction. Accessed February 5, 2024, https://www.nyc.gov/html/dot/html/infrastructure/construction.shtml

24. New York City Department of Transportation. Street Design Manual: Capital Projects. Accessed February 5, 2024, https://www.nycstreetdesign.info/process/capital-projects

25. Goin DE, Riddell CA. Comparing two-way fixed effects and new estimators for difference-in-differences: A simulation study and empirical example. Epidemiology. 2023;34(4)doi:10.1097/EDE.0000000000001611

26. Goodman-Bacon A. Difference-in-differences with variation in treatment timing. J Econom. 2021;225(2):254–277. doi:10.1016/j.jeconom.2021.03.014

27. Callaway B, Sant’Anna PHC. Difference-in-Differences with multiple time periods. J Econom. 2021;225(2):200–230. doi:10.1016/j.jeconom.2020.12.001

28. Branas CC, Kondo MC, Murphy SM, South EC, Polsky D, MacDonald JM. Urban blight remediation as a cost-beneficial solution to firearm violence. Am J Public Health. 2016;106(12):2158–2164. doi:10.2105/ajph.2016.303434

29. New York City Department of Transportation. Protected Streets. Accessed February 13, 2024, https://www.nyc.gov/html/dot/html/infrastructure/protectedst.shtml

30. New York City Department of Transportation. Street Works Manual. Accessed February 14, 2023, https://streetworksmanual.nyc/

